# Fertility rates across generations in twins and singletons: A total population study in Finland

**DOI:** 10.64898/2026.05.20.26353670

**Authors:** Sarah Niemi de Paiva, Mikaela Hukkanen, Antti Latvala, Jaakko Kaprio, Stephanie Zellers

## Abstract

**Title:** Fertility Rates Across Generations in Monozygotic Twins, Dizygotic Twins, and Non-Twin Controls in the Finnish Twin Cohort

**Study question:** Does twin status and zygosity (monozygotic vs. dizygotic; same-sex vs. opposite-sex) predict fertility outcomes and intergenerational reproductive patterns compared with singletons?

**Summary answer:** Among females, dizygotic twins had modestly higher completed fertility than singletons and monozygotic twins and were more likely to have a twin birth. Fertility did not differ meaningfully among males. These differences were restricted to the twin generation and did not persist in the next generation, indicating sex-specific and generation-specific effects rather than intergenerational transmission.

**What is known already:** Dizygotic twinning is associated with heritable hyperovulation and higher natural fertility but less is known about whether being a twin or zygosity influences reproductive outcomes across generations.

**Study design, size, duration:** A population-based longitudinal cohort study using part of the Finnish Twin Cohort and national population registers. Participants included monozygotic (MZ; N = 4,068), same-sex dizygotic (SSDZ; N = 8,890), opposite-sex dizygotic (OSDZ; N = 8,474) twins, and singleton controls (N = 1,193,404) born between 1945-1957 (total N =1,254,103; 49.1% female), their mothers, their children, and their grandchildren.

**Participants/materials, setting, methods:** Fertility outcomes (number of biological children, age at first birth, childlessness, multiple births) were derived from Finnish population registers. Analyses followed a preregistered plan (https://osf.io/qbwv3)

**Main results and the role of chance:** Differences in fertility between singletons and twins were modest and varied by sex and zygosity. Differences were observed generally in the mothers of twins and female twins themselves, with limited differences in the offspring of twins as compared to the offspring of singletons. Twins were slightly older at first birth, had fewer total biological offspring, but were more likely to have a twin birth. Dizygotic twins in particular differed from monozygotic twins and singletons.

**Limitations, reasons for caution:** Findings are limited to individuals born in mid-20th-century Finland and thus generalizability to recent populations or non-Nordic contexts may be restricted. Further, analyses are observational, and causal inference is limited due to alternative motivation behind fertility rates like social or cultural reasons.

**Wider implications of the findings:** These findings suggest that zygosity and sex interact to shape reproductive outcomes, offering insight into genetic and environmental contributions to fertility. They highlight the value of large twin cohorts for studying intergenerational reproductive trends and the representativeness of twins in population-based fertility research.

**Study funding/competing interest(s):** As a supporting activity to ongoing research projects, funding for this project was provided by the Broad Trauma Initiative at the Broad Institute of MIT and Harvard.

The authors declare no conflict of interest.

---

Human fertility rate in demography is quantified through an individual’s number of biological children (Skakkebæk *et al*., 2022). Understanding fertility patterns has become increasingly important as population reproductive trends shift toward delayed childbearing, smaller family sizes, and an increased reliance on assisted reproductive technologies. These changes carry significant implications for the prevalence of multiple births, as parity, maternal age and fertility treatments are primary drivers of twinning (Hoekstra *et al*., 2008). Understanding the genetic, physiological, and social factors (Rodgers et al., 2001, Hart, 2016, Colleran, 2016) that shape human fertility at an individual and population level is essential for ensuring that healthcare systems are structured to support the unique physiological and social demands of a changing demographic.

Understanding how twin status and zygosity can be both an outcome of and influence on fertility could inform reproductive health patterns in contemporary populations and can also inform our understanding of the genetic and physiological mechanisms underlying human reproductionAn important methodological consideration, however, is the extent to which findings from twins can be generalized to the broader population of singletons. Differences between multiple and single births on factors related to prenatal development or birth conditions may influence later reproductive outcomes. Thus, fertility represents a domain where subtle but meaningful differences may exist, making it an important outcome for testing biological hypotheses or the generalizability of twin-based research.

Previous research has investigated whether twins exhibit different fertility rates compared to singletons. Some studies report that female twins, particularly those with a male co-twin, may experience reduced lifetime fertility due to prenatal hormonal influences and developmental constraints (Bütikofer *et al*., 2019), but other research argues that twins’ fertility patterns overall align with those of the general population (Kohler et al., 2002).

One factor that could influence fertility in twins is the increased likelihood of dizygotic [DZ] twins having twin births themselves (Lichtenstein *et al*., 1996). Monozygotic [MZ] twins result from the division of a single fertilized egg whereas DZ twins arise from the fertilization of two separate ova (Smith and Penrose, 1955). MZ twinning is largely hypothesized to arise by random chance (van Dongen *et al*., 2024) it is not familial nor are there known environmental determinants. In contrast the tendency for DZ twinning is heritable, meaning that female DZ twins have a higher probability of conceiving multiples themselves compared to singletons (van Dongen *et al*., 2024).

The increased likelihood of DZ twins conceiving twins is thought to result from inherited genetic variants that promote hyperovulation where elevated follicle-stimulating hormone levels or heightened ovarian sensitivity lead to the release of multiple ova during a single menstrual cycle, thus increasing the probability of twin conception (Hoekstra *et al*., 2008). However, an opposing biological mechanism could limit later life fertility of female twins gestated with a male co-twin (Lummaa *et al*., 2007). In opposite-sex DZ [OSDZ] twin pairs, prenatal exposure to sex-specific hormonal environments could introduce asymmetric developmental effects, potentially constraining later reproductive outcomes among female twins (Lummaa *et al*., 2007). Evidence for this phenomenon is mixed, as studies have found no differences between same-sex DZ [SSDZ] females compared to OSDZ females in fertility and reproductive cancers (Ahrenfeldt *et al*., 2015; Rose *et al*., 2002).

These competing biological mechanisms, developmental constraints versus genetically mediated hyperovulation, operate in opposite directions. As a result, any fertility differences associated with twin status are expected to be modest, sex-specific, and dependent on zygosity. This genetic predisposition to having a twin pregnancy suggests that dizygotic twins, particularly females, may have different fertility patterns than monozygotic twins or singletons (Lichtenstein *et al*., 1996). Findings from historical data, however, emphasize the importance of environmental and social factors in shaping fertility outcomes, rather than just considering twin status. Research on pre-industrial populations in Europe have demonstrated that reproduction rates are influenced by premature mortality rates and socioeconomic factors, which may differentially affect the fertility of twins compared to singletons (Rickard *et al*., 2022).

For example, in studies based on pre-industrialized populations, twin pregnancies were even more high risk than at present and more often resulted in the death of the mother and/or the twins than singleton births. These birth complications may have counterbalanced the possible genetic liability for higher fertility in twin mothers, often also seen in older mothers with multiple prior offspring. In more recent samples, mothers of naturally conceived DZ twins are taller and have higher BMI as compared to mothers of naturally conceived MZ twins or mothers of DZ twins conceived with assistive reproductive technology (Hoekstra *et al*., 2010; Hubers *et al*., 2024). These differences in body composition may reflect greater capacity to carry a twin pregnancy to term as well as higher hormone levels, including levels of follicle-stimulating hormone (Hubers *et al*., 2024). Indeed, genes associated with twinning are associated with biology and body size (Mbarek *et al*., 2023).

Furthermore, the timing of reproduction also contributes to fertility outcomes as younger parents might be less resilient to parenting-related physical, emotional, and economic stress, and also experience a higher allostatic load overall (Barban, 2013; Grundy and Read, 2015), while on the other hand early reproduction can lead to shorter generation times, thus promoting the overall reproductive success of a family line. In contrast, the likelihood of dizygotic twinning also increases with maternal age (Hubers *et al*., 2024; Otta *et al*., 2016). This intricate relationship between twinning, maternal age, and fertility outcomes has emerged as a crucial area of contemporary demographic research given the widespread trend of delayed reproduction in modern populations.

Despite substantial progress in twin and fertility research, several gaps in the literature remain. Many earlier studies are limited to a single generation, which constrains the ability to detect intergenerational reproductive patterns. Further, there is limited previous research that has differentiated between SSDZ and OSDZ twins. The present study addresses these gaps by leveraging the Finnish Twin Cohort and linked national registers to examine fertility outcomes across multiple generations in a large, fully population-based sample.

This study utilized data from the Finnish Twin Cohort [FTC] linked with national population registers as part of the TwinRegistry project. This data allows for direct comparison of fertility through a large, population-based reference sample of all singletons born in the same years as part of the FTC sample (Zellers *et al*., 2025). The dataset also enables multi-generational analyses, by linking twins and the reference population to their own parents and their children and grandchildren. Prior assessments of the FTC indicate that twins closely resemble their birth cohort on demographic indicators such as education, mortality, employment, residence, and marital status (Zellers *et al*., 2025).

First, we hypothesized that dizygotic twins would differ in fertility rates compared to monozygotic twins and singletons (H1). Specifically, we expected female DZ twins to have higher lifetime fertility than female MZ twins and singletons. However, we anticipated these differences to be modest in size. Second, we hypothesized that the fertility rates of the children of twins would vary by their parents’ zygosity (H2). We expected that female children of DZ twins would show higher fertility rates than female children of MZ twins, consistent with the heritable nature of DZ twinning. Finally, we hypothesized that the fertility rates of OSDZ twins would differ from those of same-sex twins (MZ and SSDZ; H3). Together, these hypotheses test whether twin zygosity and sex predict fertility patterns across generations. All analyses followed a preregistered analysis plan to ensure transparency and reproducibility (OSF preregistration: https://osf.io/qbwv3).

## Methods

### Sample

The Finnish TwinRegistry is a registry-based data resource that integrates questionnaire data from the FTC with national administrative records (Zellers *et al*., 2025). The FTC was initially established in 1974 and includes twins from same-sex pairs born in Finland before 1958 (Kaprio *et al*., 2019). TwinRegistry expands the FTC by linking twin participants to their family members, including spouses, siblings, parents and children, and to national health, social and economic registers curated by Statistics Finland. The dataset also includes a reference population of all Finnish individuals born between 1945 and 1957 and their linked relatives, allowing for analyses of generalizability between twins and singletons. Through the registry we additionally have identified opposite-sex pairs.

We conducted a retrospective cohort study using TwinRegistry with the focus population being twins born between 1945 and 1957. The approximately 1.25 million non-twin individuals born in Finland during the same period were included for comparison. Family linkages were drawn for both twins and non-twin controls amounting to 4.6 million unique participants spanning multiple generations. See Figure 1 for an illustration of the sample structure. All aspects of this research complied with the regulations governing the use of administrative data in Finland. Follow up ended on December 31, 2020, for the present dataset.

**Figure 1.**
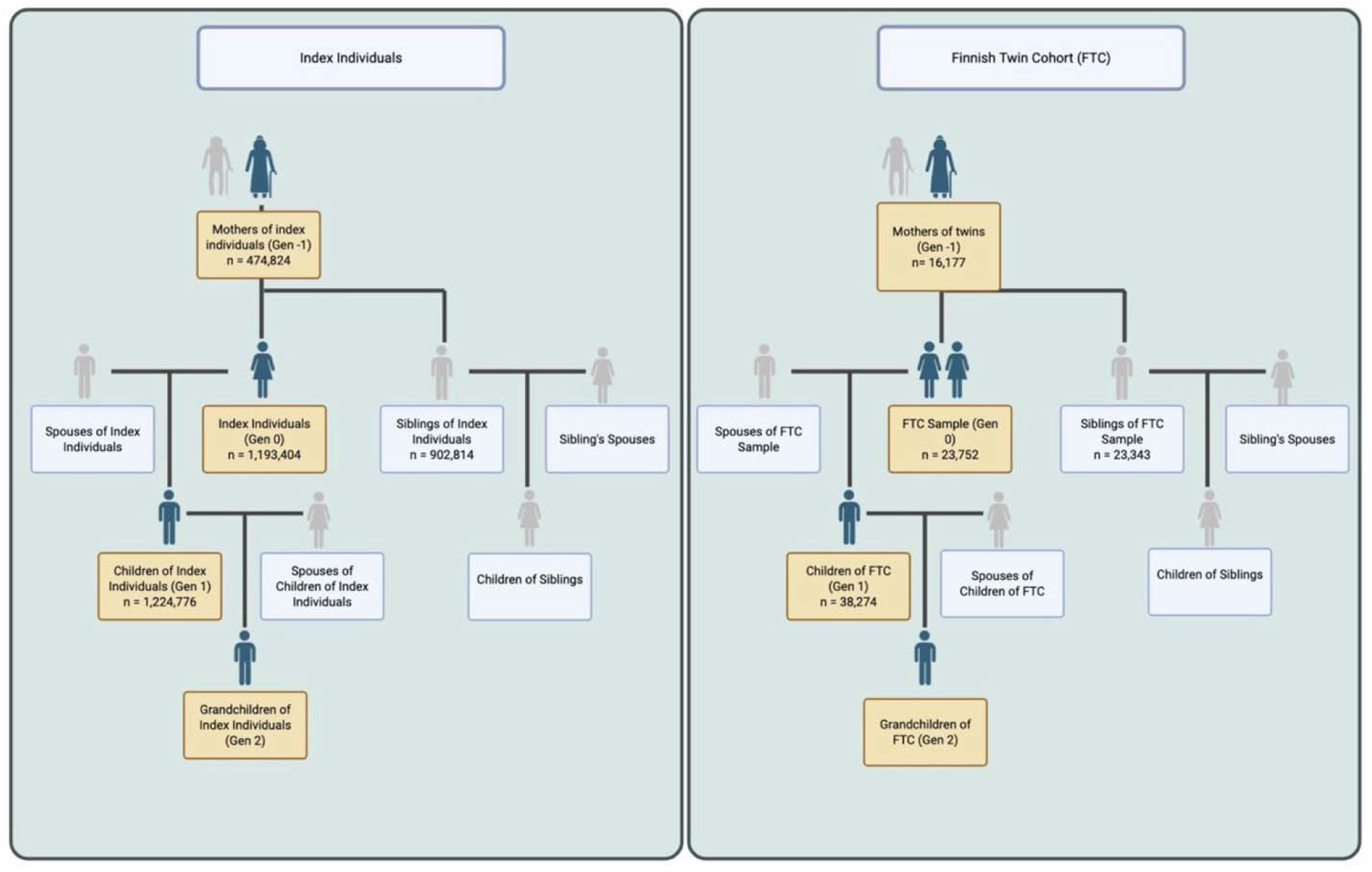
Caption: Schematic overview of the linked register data used in the study, combining the Finnish population birth cohort (1945–1957) with the Finnish Twin Cohort. Figure 1 Alt Text: A diagram in the style of a family tree illustrating the linked register data used in the study.

The base analytic sample (Generation 0) included 1,277,849 individuals with complete fertility data: non-twin controls (n = 1,254,103), SSDZ twins (n = 8,890), OSDZ twins (n = 8,474), MZ twins (n = 4,068), and twins of unknown zygosity (XZ; n = 2,314). Complete multigenerational family linkages were available for both twins and controls, allowing direct comparison of fertility patterns across generations (Figure 1). We describe the mothers of the twin and singleton core sample as Generation −1, comprising 474,824 mothers of singletons and 16,177 mothers of multiples. It is important to acknowledge selection bias here, in that the mothers of multiples includes all mothers of FTC participants, as well as mothers of singleton controls who have had a multiple birth that was not included in the FTC (birth before or after the window 1945-1957). In general, there is selection bias because Generation -1 individuals are included on the condition of having at least one child.

The offspring of the twin/singleton core sample are described as Generation 1 and comprised 1,224,776 children of singletons and 38,274 children of twins. The offspring of Generation 1 are described as Generation 2 (the grandchildren of the Generation 0 twin/singleton core sample). Data quality checks were performed, and records exhibiting inconsistencies in family structure or reproductive timelines were identified and resolved.

### Measures

#### Predictors

The primary predictors were twin status and zygosity. Twin status was coded as twin or singleton. Zygosity for twins was classified as MZ, SSDZ, OSDZ, or XZ and these twin zygosity groups were all compared against singletons. Twins from same-sex pairs were originally identified from birth records and contacted for participation in FTC questionnaires. Zygosity classification relied on validated questionnaire-based assessments collected surveys in 1975 and 1981 and supplemented by genotyping in later DNA collections according to established Finnish Twin Cohort procedures (Kaprio *et al*., 2019; Sarna *et al*., 1978). The XZ group consists either of twins identified from birth records who did not respond to any questionnaires and may therefore differ systematically from classified twins due to nonresponse bias, or of twins who replied to the questionnaires but whose responses were inconsistent on the key items used to classify zygosity. Thus, the XZ group contains SSDZ and MZ twins. All opposite-sex twins could be comprehensively identified as born on the same day to the same mother but being of opposite sex in the population register. .

Intergenerationally, twin status and zygosity were coded centered on Generation 0. For example, mothers of twins and singletons in Generation -1 would be grouped as “twin mothers” or “singleton mothers” based on their offspring in Generation 0. We also examined siblings of Generation 0 singletons to identify any multiple births that were not included in the Generation 0 sample. In other words, a Generation -1 mother of a Generation 0 singleton may have had a multiple birth outside of the years 1945-1957. Similarly, the offspring Generations 1 and 2 were coded based on their parents/grandparents respectively in Generation 0.

#### Outcomes

Fertility outcomes were derived from verified parent–child linkages in national population registers and reconstructed as longitudinal reproductive histories across generations. Primary outcomes included the total number of biological children, age at first live birth computed from the birth date of the parent and offspring, a binary indicator for childlessness (zero biological children as compared to any biological children), a binary indicator for whether the individual had any multiple birth (twin or higher order multiple) or only had single births.

This indicator for multiple births could only be calculated for Generation 0, based on availability of both familial linkage data and exact birthdates, allowing for identification of multiple births. Exact birthdates are not available for Generation 2, making it impossible to identify multiple births by Generation 1 individuals. All outcomes were based on register-verified biological relationships and were used in both descriptive summaries and inferential mixed-effects regression models.

### Statistical Analysis

#### Descriptive Statistics

We generated descriptive tables of sample sizes, birth years, and fertility outcomes for each generation (Generation -1, Generation 0, and Generation 1, Generation 2) stratified by sex and twin status/zygosity. Group-specific means/medians and standard deviations were calculated for continuous outcomes and group-specific frequencies for binary outcomes to provide a baseline characterization of fertility patterns across groups and generations.

#### Mixed-Effects Models

To formally test associations between twin status (twin or singleton), zygosity (MZ, SSDZ, OSDZ, XZ, or singleton), and fertility outcomes, we employed mixed-effects regression models estimated using the lme4 package in RStudio (Bates *et al*., 2015; R Core Team, 2022). All models included a random intercept for family ID to account for clustering within families. All analyses were conducted separately by parental generation (−1, 0, 1) and sex to capture potential differences in fertility patterns from either biological (transmission of heritable variation) or sociocultural sources. In models with twin status as the predictor, the reference category was singleton. In models with zygosity as the predictor, the reference category was also singleton.

Model type varied by outcome, as there are different distributions and assumptions based on variable type. For number of children (count variable) we utilized Poisson mixed-effects regression models (function glmer with family = Poisson). For age at first birth (continuous variable) we used linear mixed-effects regression models (function lmer). Lastly for childlessness and twin births (binary variables) we used binomial mixed-effects regression models (function glmer with family = binomial).

#### Supplemental Analyses

For Generation 1, as some individuals were not old enough to have completed lifetime fertility by the end of follow up, we conducted analyses two ways. We ran all models with all Generation 1 individuals, regardless of birth year (results presented in supplement). We additionally ran all models with only the subset of Generation 1 individuals who were at least 40 years old by the end of follow up (results presented in main text). In other words, the restricted sample consisted of Generation 1 individuals born by December 31, 1980 (N=682,073; 54% of total Generation 1 sample). This choice was made to assess the influence of the proportion of censored observations in our results, and we present both sets of results for transparency.

For the primary models, singletons were the reference category for all comparisons. To further understand differences between zygosity groups, we additionally ran twin-only models with MZ as the reference category. We chose MZ as the reference category because it is hypothesized that the genetic component of twinning only increases likelihood of DZ twinning, whereas MZ twinning is akin to random chance (van Dongen *et al*., 2024). We thus were interested in investigating whether MZ twins differed from DZ and XZ groups. Results are presented in the supplement.

## Results

### Fertility characteristics across the study sample

Table 1 presents descriptive fertility characteristics for Generation −1, comprising the mothers of singleton controls and twins in the index population. Table 2 presents the same descriptive statistics for Generation 0, the twins and singletons born 1945-1957. Lastly Table 3 (subset of sample born by 1980) and Supplemental Table 1 (full sample) present descriptive statistics for Generation 1, the offspring of the twins and singletons. All descriptive tables are stratified by sex, twin status, and zygosity. Patterns of group differences observed in these tables were tested in the mixed effects models. Sex distributions were approximately balanced across groups, except among XZ twins, where 58.7% were male.

**Table 1:** Descriptives on Fertility of Mothers of Finnish Twin Cohort and Singletons (Generation -1)

|  |  |  |  |
| --- | --- | --- | --- |
| **Mothers Stratified by Twin Status** | **N** | **Mean Age at First Birth (SD)** | **Mean Number of Children (SD)** |
| Mothers of Singletons | 474,824 | 27.8 (6.41) | 2.7 (1.6) |
| Mothers of Multiples | 16,177 | 27.2 (5.78) | 4.5 (2.0) |
| **Mothers Stratified by Offspring Zygosity** |  |  |  |
| MZ | 1,203 | 28.2 (6.44) | 3.67 (1.68) |
| SSDZ | 2,644 | 29.5 (6.08) | 3.66 (1.71) |
| OSDZ | 2,866 | 28.5 (5.69) | 3.88 (1.82) |
| XZ | 1,155 | 27.5 (6.14) | 4.17 (1.96) |
| Twins/Multiples Not in FTC* | 8,309 | 25.8 (5.17) | 5.14 (1.97) |

\*Twins/Multiples Not in FTC: mothers who had a multiple birth elsewhere in the family, not included in the FTC because the birth occurred after 1958.
Note: To be included in each group, mothers were required to have a minimum number of children: at least 1 for the “Mothers of Singletons” group, at least 2 for the “Mothers of Multiples” group, and at least 3 for the “Twins/Multiples Not in FTC” group (the singleton included in the study plus a multiple birth among the singleton’s siblings). MZ - monozygotic, SSDZ - same sex dizygotic, OSDZ - opposite sex dizygotic, XZ - unknown zygosity

**Table 2:** Descriptives on Fertility of Twins and Singletons (Generation 0)

|  |  |  |  |  |  |  |  |  |  |  |
| --- | --- | --- | --- | --- | --- | --- | --- | --- | --- | --- |
|  | **N** |  | **Mean Age at First Birth (SD)** |  | **% Childless** |  | **Mean Number of Children (SD)** |  | **% Twin Births** |  |
| **Twin Status** | **M** | **F** | **M** | **F** | **M** | **F** | **M** | **F** | **M** | **F** |
| Singletons | 607,570 | 585,834 | 27.2<br>(5.4) | 24.7<br>(4.9) | 24.5% | 17.8% | 1.70<br>(1.41) | 1.80<br>(1.32) | 1.84% | 1.87% |
| Twins | 12,064 | 11,688 | 27.4<br>(5.3) | 25.2<br>(5.1) | 28.9% | 20.4% | 1.61<br>(1.43) | 1.75<br>(1.36) | 1.77% | 2.28% |
| **Zygoty Stratified** |  |  |  |  |  |  |  |  |  |  |
| MZ | 1,906 | 2,162 | 27.2<br>(5.3) | 25.5<br>(5.1) | 26.4% | 22.4% | 1.66<br>(1.41) | 1.68<br>(1.33) | 1.68% | 1.71% |
| SSDZ | 4,558 | 4,332 | 27.4<br>(5.3) | 25.4<br>(5.2) | 26.5% | 19.6% | 1.66<br>(1.37) | 1.76<br>(1.36) | 1.80% | 2.29% |
| OSDZ | 4,239 | 4,235 | 27.4<br>(5.3) | 25.1<br>(4.9) | 27.6% | 17.3% | 1.68<br>(1.49) | 1.86<br>(1.36) | 1.84% | 2.67% |
| XZ | 1,390 | 960 | 27.4<br>(5.5) | 24.7<br>(4.9) | 44.6% | 33.6% | 1.19<br>(1.38) | 1.41<br>(1.34) | 1.54% | 1.77% |

Note: M - males, F - females, MZ - monozygotic, SSDZ - same sex dizygotic, OSDZ - opposite sex dizygotic, XZ - unknown zygoty

**Table 3:** Descriptives on Fertility of Children of Twins and Children of Singletons (Generation 1)

|  |  |  |  |  |  |  |  |  |
| --- | --- | --- | --- | --- | --- | --- | --- | --- |
|  | **N Born By 1980** |  | **Mean Age at First Birth (SD)** |  | **% Childless** |  | **Mean N Children (SD)** |  |
| **Twin Status** | **M** | **F** | **M** | **F** | **M** | **F** | **M** | **F** |
| Singletons | 338,467 | 323,775 | 29.6<br>(5.5) | 27.6<br>(5.4) | 28.7% | 19.7% | 1.63<br>(1.45) | 1.87<br>(1.42) |
| Twins | 9,964 | 9,687 | 29.7<br>(5.5) | 27.5<br>(5.4) | 27.6% | 19.7% | 1.67<br>(1.47) | 1.90<br>(1.45) |
| **Zygoty Stratified** |  |  |  |  |  |  |  |  |
| MZ | 1,760 | 1,650 | 29.7<br>(5.6) | 27.3<br>(5.4) | 29.0% | 19.6% | 1.63<br>(1.42) | 1.90<br>(1.49) |
| SSDZ | 3,902 | 3,853 | 29.7<br>(5.5) | 27.6<br>(5.4) | 27.7% | 19.7% | 1.65<br>(1.47) | 1.90<br>(1.45) |
| OSDZ | 3,616 | 3,486 | 29.7<br>(5.5) | 27.6<br>(5.3) | 26.7% | 19.7% | 1.70<br>(1.51) | 1.91<br>(1.43) |
| XZ | 686 | 698 | 29.3<br>(5.5) | 26.7<br>(5.6) | 29.2% | 19.8% | 1.64<br>(1.46) | 1.91<br>(1.46) |

Note: M - males, F - females, MZ - monozygotic, SSDZ - same sex dizygotic, OSDZ - opposite sex dizygotic, XZ - unknown zygoty. This table includes only individuals born by 1980 to ensure completed fertility. Table including all individuals, regardless of birth year, presented in the supplement.

### Associations between twin status, zygosity and reproductive outcomes

#### Generation -1

Mixed effect model results are presented in Tables 4 and 5. Note the outcome “childless” and “twin birth” are not observed in Generation -1 as those are criteria for inclusion in the sample (i.e. all individuals in the Generation -1 sample have at least one child; having a twin birth is the basis on which Generation -1 mothers are grouped). In Generation −1 females, twin motherhood was associated with an earlier age at first birth (β = -0.61) and substantially higher completed fertility relative to mothers of singletons (IRR = 1.67). Fertility differences varied by offspring zygosity, with the largest effects observed among mothers with multiple births outside the FTC (age at first birth β = -1.98; number of biological children IRR = 1.91), which logically follows that this group of mothers have both a singleton birth between 1945-1957 and a multiple birth during some year outside that range. In contrast, mothers of FTC MZ, SSDZ, and OSDZ twins were on average slightly older at first birth as compared to mothers of singletons (MZ β = 0.43, OSDZ β = 0.68, SSDZ β =1.69). These results reflect the defining feature of this generation—selection into motherhood—and primarily serve as contextual background for subsequent generational analyses.

**Table 4:**
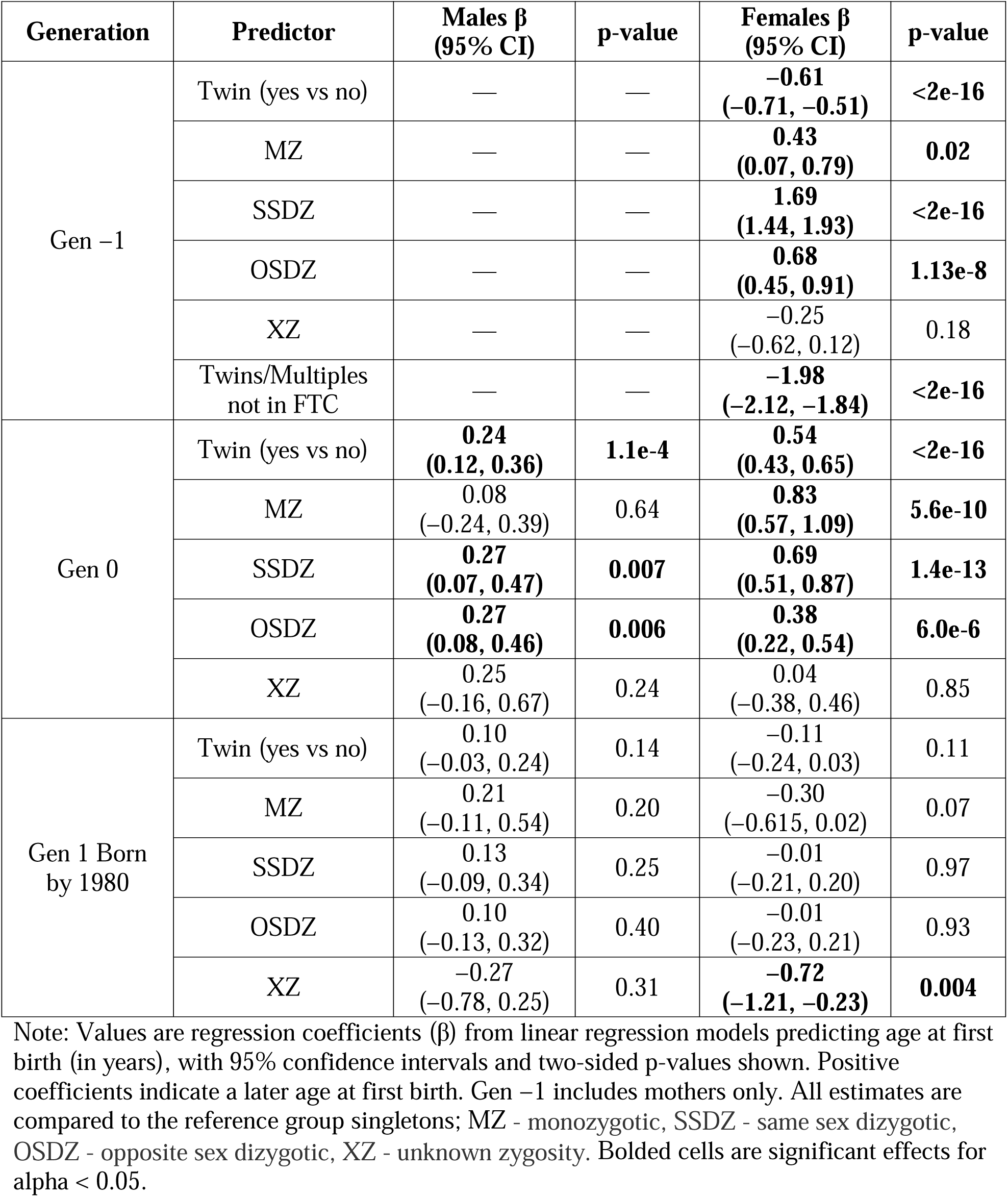
Twin vs. Singleton Differences in Age at First Birth.

| **Generation** | **Predictor** | **Males $\beta$**<br>**(95% CI)** | **p-value** | **Females $\beta$**<br>**(95% CI)** | **p-value** |
| --- | --- | --- | --- | --- | --- |
| Gen -1 | Twin (yes vs no) | — | — | **-0.61**<br>(-0.71, -0.51) | **<2e-16** |
|  | MZ | — | — | **0.43**<br>(0.07, 0.79) | **0.02** |
|  | SSDZ | — | — | **1.69**<br>(1.44, 1.93) | **<2e-16** |
|  | OSDZ | — | — | **0.68**<br>(0.45, 0.91) | **1.13e-8** |
|  | XZ | — | — | -0.25<br>(-0.62, 0.12) | 0.18 |
|  | Twins/Multiples<br>not in FTC | — | — | **-1.98**<br>(-2.12, -1.84) | **<2e-16** |
| Gen 0 | Twin (yes vs no) | **0.24**<br>(0.12, 0.36) | **1.1e-4** | **0.54**<br>(0.43, 0.65) | **<2e-16** |
|  | MZ | 0.08<br>(-0.24, 0.39) | 0.64 | **0.83**<br>(0.57, 1.09) | **5.6e-10** |
|  | SSDZ | **0.27**<br>(0.07, 0.47) | **0.007** | **0.69**<br>(0.51, 0.87) | **1.4e-13** |
|  | OSDZ | **0.27**<br>(0.08, 0.46) | **0.006** | **0.38**<br>(0.22, 0.54) | **6.0e-6** |
|  | XZ | 0.25<br>(-0.16, 0.67) | 0.24 | 0.04<br>(-0.38, 0.46) | 0.85 |
| Gen 1 Born<br>by 1980 | Twin (yes vs no) | 0.10<br>(-0.03, 0.24) | 0.14 | -0.11<br>(-0.24, 0.03) | 0.11 |
|  | MZ | 0.21<br>(-0.11, 0.54) | 0.20 | -0.30<br>(-0.615, 0.02) | 0.07 |
|  | SSDZ | 0.13<br>(-0.09, 0.34) | 0.25 | -0.01<br>(-0.21, 0.20) | 0.97 |
|  | OSDZ | 0.10<br>(-0.13, 0.32) | 0.40 | -0.01<br>(-0.23, 0.21) | 0.93 |
|  | XZ | -0.27<br>(-0.78, 0.25) | 0.31 | **-0.72**<br>(-1.21, -0.23) | **0.004** |

Note: Values are regression coefficients ( $\beta$ ) from linear regression models predicting age at first birth (in years), with 95% confidence intervals and two-sided p-values shown. Positive coefficients indicate a later age at first birth. Gen -1 includes mothers only. All estimates are compared to the reference group singletons; MZ - monozygotic, SSDZ - same sex dizygotic, OSDZ - opposite sex dizygotic, XZ - unknown zygosity. Bolded cells are significant effects for alpha < 0.05.

**Table 5:** Twin vs. Singleton Differences in Total Number of Biological Children.

|  |  |  |  |  |  |
| --- | --- | --- | --- | --- | --- |
| **Generation** | **Predictor** | **Males IRR**<br>**(95% CI)** | **p-value** | **Females IRR**<br>**(95% CI)** | **p-value** |
| Gen -1 | Twin (yes vs no) | — | — | 1.67<br>(1.66, 1.68) | <2e-16 |
|  | MZ | — | — | 1.36<br>(1.32, 1.40) | <2e-16 |
|  | SSDZ | — | — | 1.36<br>(1.33, 1.38) | <2e-16 |
|  | OSDZ | — | — | 1.44<br>(1.41, 1.47) | <2e-16 |
|  | XZ | — | — | 1.55<br>(1.50, 1.59) | <2e-16 |
|  | Twins/Multiples<br>not in FTC | — | — | 1.91<br>(1.89, 1.93) | <2e-16 |
| Gen 0 | Twin (yes vs no) | **0.95**<br>**(0.93, 0.96)** | **4.6e-11** | **0.97**<br>**(0.96, 0.98)** | **2.0e-5** |
|  | MZ | 0.97<br>(0.93, 1.01) | 0.14 | **0.93**<br>**(0.90, 0.96)** | **9.8e-6** |
|  | SSDZ | **0.97**<br>**(0.95, 0.99)** | **0.04** | **0.97**<br>**(0.95, 0.99)** | **0.02** |
|  | OSDZ | 0.99<br>(0.96, 1.01) | 0.28 | **1.03**<br>**(1.01, 1.05)** | **0.01** |
|  | XZ | 0.69<br>(0.66, 0.73) | **<2e-16** | **0.78**<br>**(0.74, 0.83)** | **<2e-16** |
| Gen 1 Born by<br>1980 | Twin (yes vs no) | 1.02<br>(0.99, 1.04) | 0.13 | 1.02<br>(0.99, 1.03) | 0.08 |
|  | MZ | 0.99<br>(0.94, 1.04) | 0.63 | 1.02<br>(0.98, 1.06) | 0.34 |
|  | SSDZ | 1.01<br>(0.98, 1.04) | 0.70 | 1.012<br>(0.99, 1.04) | 0.37 |
|  | OSDZ | **1.04**<br>**(1.01, 1.07)** | **0.02** | 1.018<br>(0.99, 1.05) | 0.20 |
|  | XZ | 1.01<br>(0.94, 1.08) | 0.83 | 1.007<br>(0.95, 1.07) | 0.81 |

Note: Values are incidence rate ratios (IRRs = $\exp(\beta)$ ) from Poisson regression models predicting the number of children, with 95% confidence intervals and two-sided p-values shown. IRRs greater than 1 indicate higher completed fertility. Gen -1 includes mothers only. All estimates are compared to the reference group singletons; MZ - monozygotic, SSDZ - same sex dizygotic, OSDZ - opposite sex dizygotic, XZ - unknown zygosity. Bolded cells are significant effects for $\alpha < 0.05$ .

#### Generation 0

Results from mixed-effects regression models predicting age at first birth and number of children are shown in Tables 4 and 5 and summarized visually in Figure 2A. In Generation 0, being a twin was associated with a slightly later age at first birth for both males (β = 0.24) and females (β = 0.54). Zygosity-specific models indicated later age at first birth among female twins of known zygosity (MZ β = 0.83, SSDZ β =0.69, and OSDZ β = 0.38) relative to singletons.

**Figure 2.**
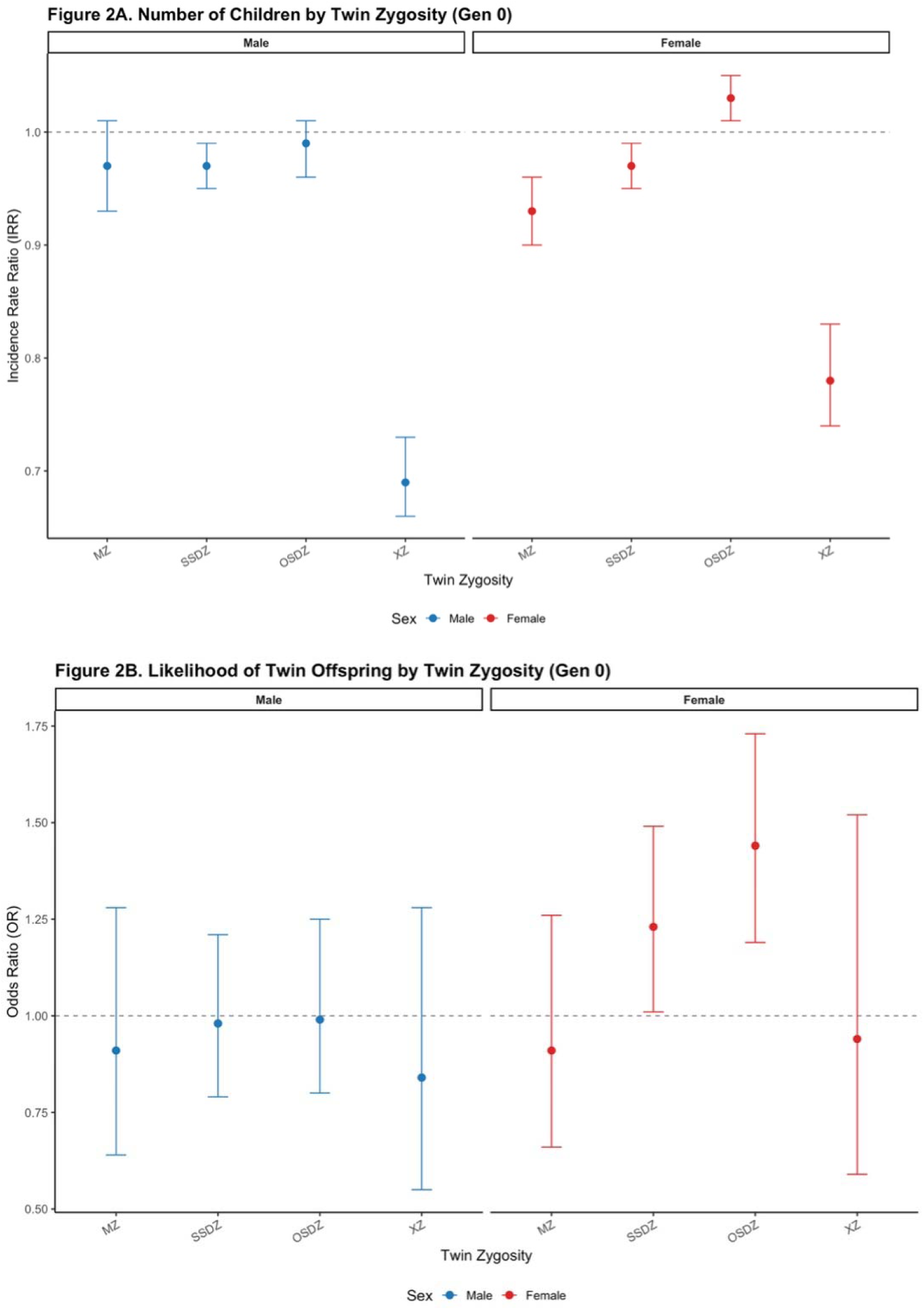
Caption: Association between twin zygosity and reproductive outcomes in Generation 0. Panel A shows incidence rate ratios (IRRs) from count models estimating the number of biological children, with singletons as the reference group. Panel B shows odds ratios (ORs) from logistic regression models estimating the likelihood of having twin offspring, also with singletons as the reference group. Estimates are shown separately for males and females in faceted panels (males left, females right). Points represent model estimates and horizontal lines indicate 95% confidence intervals; dashed horizontal lines mark the null value (IRR or OR = 1). Figure 2A Alt Text: Line graphical representations of Poisson regression model results for number of children by zygosity, there are two panels in the figure to show results for males and females separately. Figure 2B Alt Text: Line graphical representations logistic regression model results for likelihood of twin births by zygosity, there are two panels in the figure to show results for males and females separately.

Male SSDZ (β = 0.27) and OSDZ (β = 0.27) twins similarly were slightly older than singletons at their first birth.

Regarding completed fertility (Figure 2A), twins had modestly fewer biological children than singletons overall (IRR = 0.95 for males; IRR = 0.97 for females). Zygosity-specific contrasts showed that female OSDZ twins had modestly higher fertility than singletons (IRR = 1.03), whereas female MZ, SSDZ, and XZ twins had modestly lower fertility (MZ IRR = 0.93, SSDZ IRR = 0.97, XZ IRR = 0.78). Among males, XZ twins had less offspring than singletons (IRR = 0.69), driving the observed twin-singleton differences.

With respect to childlessness (Table 6), male twins were more likely to be childless as compared to male singletons (OR=1.26) but we observed no overall effect of being a twin for females (OR=0.69). When comparing zygosity groups to singletons, SSDZ (OR = 1.11), OSDZ (OR = 1.17), and XZ (OR = 2.51) male twins were all more likely to be childless as compared to singletons; male MZ twins did not differ from singletons in likelihood of childlessness. Female MZ (OR = 1.33), SSDZ (OR = 1.12), and XZ (OR = 2.34) twins were similarly all more likely to be childless; female OSDZ twins did not differ from singletons on likelihood of being childless. Female Generation 0 twins were more likely to have twin offspring than singletons (Figure 2B) (OR = 1.22) and the effect was driven by dizygotic twins (both SSDZ [OR = 1.23] and OSDZ [OR = 1.44]) (Table 7). No effect was observed for males (OR = 0.96).

**Table 6:** Twin vs. Singleton Differences in Likelihood of Being Childless.

|  |  |  |  |  |  |
| --- | --- | --- | --- | --- | --- |
| **Generation** | **Predictor** | **Males OR**<br>**(95% CI)** | **p-value** | **Females OR**<br>**(95% CI)** | **p-value** |
| Gen 0 | Twin (yes vs no) | **1.26**<br>**(1.21, 1.31)** | **<2e-16** | **1.18**<br>**(1.13, 1.24)** | **3.8e-13** |
|  | MZ | 1.11<br>(1.00, 1.23) | 0.05 | **1.33**<br>**(1.20, 1.48)** | **2.7e-8** |
|  | SSDZ | **1.11**<br>**(1.04, 1.19)** | **0.002** | **1.12**<br>**(1.04, 1.21)** | **0.002** |
|  | OSDZ | **1.17**<br>**(1.10, 1.26)** | **4.1e-6** | 0.96<br>(0.89, 1.04) | 0.34 |
|  | XZ | **2.51**<br>**(2.25, 2.80)** | **<2e-16** | **2.34**<br>**(2.04, 2.68)** | **<2e-16** |
| Gen 1 Born by 1980 | Twin (yes vs no) | **0.95**<br>**(0.90, 0.99)** | **0.03** | 1.00<br>(0.94, 1.06) | 0.90 |
|  | MZ | 1.02<br>(0.92, 1.14) | 0.72 | 0.98<br>(0.85, 1.13) | 0.76 |
|  | SSDZ | 0.95<br>(0.88, 1.02) | 0.15 | 1.00<br>(0.91, 1.10) | 0.99 |
|  | OSDZ | **0.90**<br>**(0.83, 0.97)** | **0.01** | 1.00<br>(0.91, 1.10) | 0.92 |
|  | XZ | 1.03<br>(0.86, 1.23) | 0.75 | 1.02<br>(0.83, 1.27) | 0.83 |

Note: Values are odds ratios (ORs = $\exp(\beta)$ ) from logistic regression models predicting the likelihood of being childless, with 95% confidence intervals and two-sided p-values shown. ORs greater than 1 indicate higher likelihood of being childless. All estimates are compared to the reference group singletons; MZ - monozygotic, SSDZ - same sex dizygotic, OSDZ - opposite sex dizygotic, XZ - unknown zygosity. Bolded cells are significant effects for $\alpha < 0.05$ .

**Table 7:** Twin vs. Singleton Differences in Likelihood of Having Twin Offspring.

|  |  |  |  |  |  |
| --- | --- | --- | --- | --- | --- |
| **Generation** | **Predictor** | **Males OR**<br>**(95% CI)** | **p-value** | **Females OR**<br>**(95% CI)** | **p-value** |
| Gen0 | Twin (yes vs no) | 0.96<br>(0.84, 1.10) | 0.52 | **1.22**<br>**(1.08, 1.38)** | **1.4e-3** |
|  | MZ | 0.91<br>(0.64, 1.28) | 0.59 | 0.91<br>(0.66, 1.26) | 0.57 |
|  | SSDZ | 0.98<br>(0.79, 1.21) | 0.82 | **1.23**<br>**(1.01, 1.49)** | **0.04** |
|  | OSDZ | 0.99<br>(0.80, 1.25) | 0.98 | **1.44**<br>**(1.19, 1.73)** | **1.4e-4** |
|  | XZ | 0.84<br>(0.55, 1.28) | 0.41 | 0.94<br>(0.59, 1.52) | 0.81 |

Note: Values are odds ratios (ORs = $\exp(\beta)$ ) from logistic regression models predicting the likelihood of having twin offspring, with 95% confidence intervals and two-sided p-values shown. ORs greater than 1 indicate higher likelihood of having a twin birth. All estimates are compared to the reference group singletons; MZ - monozygotic, SSDZ - same sex dizygotic, OSDZ - opposite sex dizygotic, XZ - unknown zygosity. Bolded cells are significant effects for $\alpha < 0.05$ .

Taken together, these results indicate that being a twin is associated with small but systematic differences in fertility in Generation 0, with effects varying by sex and zygosity and most evident among female DZ twins.

#### Generation 1

Mixed-effects regression models for Generation 1 are reported in Tables 4 and 5 and summarized in Figure 3. Across both sexes, twin status of the parent was not associated with age at first birth (Table 4). Zygosity-specific effects were likewise modest and generally non-significant, except for daughters of XZ twins, who were slightly younger at first birth than singletons (β = -0.72).

**Figure 3.**
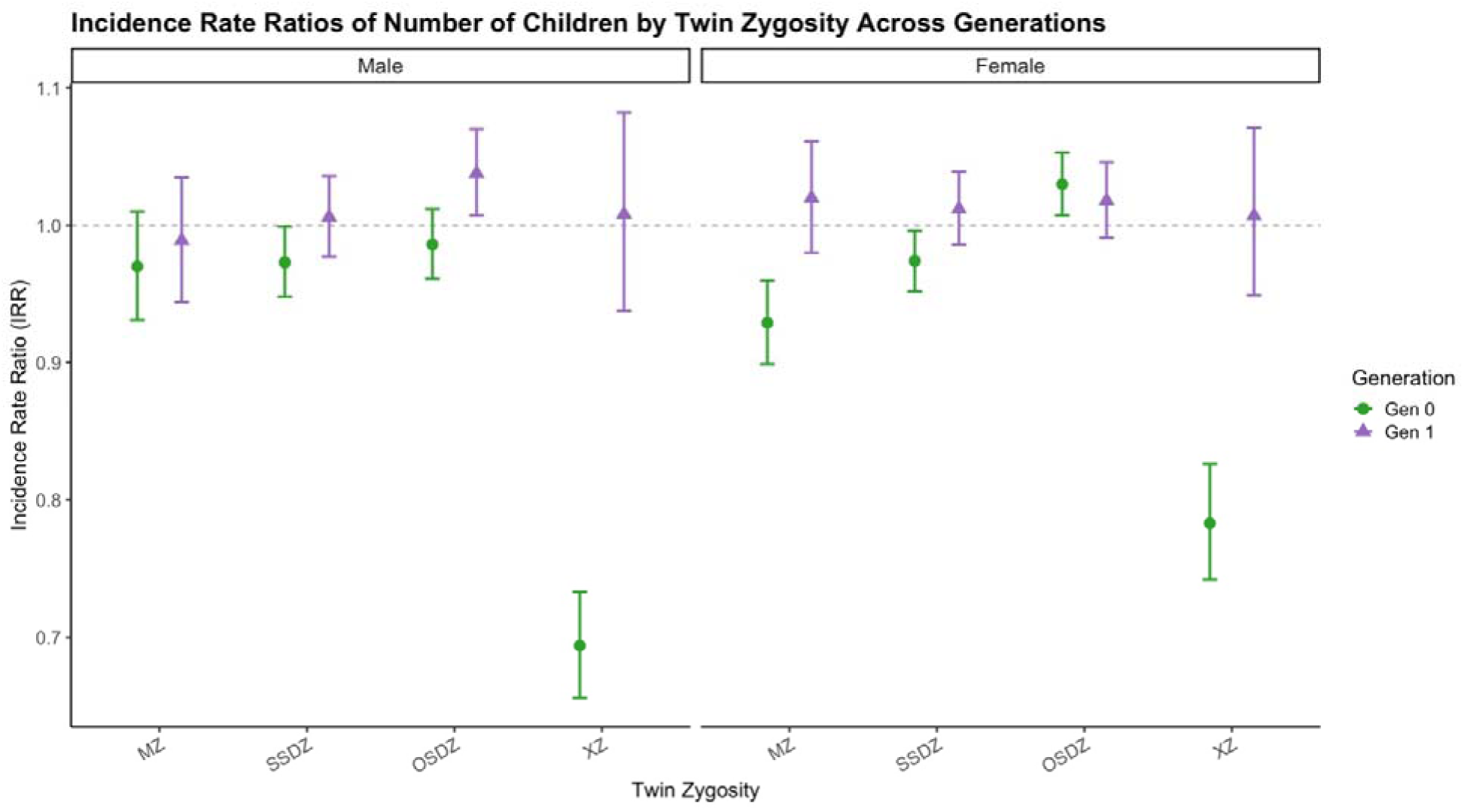
Caption: Incidence rate ratios (IRRs) and 95% confidence intervals for completed fertility by zygosity across two generations. Generation 0 estimates (green circles) reflect fertility among twins themselves, while Generation 1 estimates (purple triangles) reflect fertility among children of twins. The dashed horizontal line represents the null (IRR = 1). Figure 3 Alt Text: Line graphical representation of Poisson regression results for number of offspring by zygosity and generation, there are two panels in the figure to show results for males and females separately.

In models predicting number of biological children (Table 5), no consistent differences were observed between children of twins and children of singletons. Overall, estimates clustered closely around the null, indicating little evidence of intergenerational transmission of twin- or zygosity-related fertility differences. The same is true for likelihood of being childless (Table 6).

#### Supplemental Results Evaluating Censoring in Generation 1

When comparing the results in the full Generation 1 group to the subset born by 1980, we identified several disagreements. The results for age at first birth (Table 4 and Table S2) were in complete agreement with respect to significance and direction of effect. Within the results for number of biological children (Table 5 and Table S3), one estimate was significant in the subset born by 1980 but not in the full sample (OSDZ males having significantly more biological children than singletons). Within the results for likelihood of being childless (Table 6 and Table S4), there were the highest rate of disagreements between the full sample and the sample born by 1980, highlighting the possibility for censoring due to incomplete fertility data, as those younger individuals may not have had their first offspring by 2020, particularly in the current sociocultural trend of delayed age at first birth (Roustaei *et al*., 2019).

#### Supplemental Results Evaluating Zygosity Differences Between Twins Only

Tables S5, S6, S7, and S8 present the results for twin-only mixed effects models for each outcome, with MZ twins as the reference category, for age at first birth (S5), number of biological children (S6), likelihood of being childless (S7), and likelihood of having twin offspring (S8). We identified various differences between twin zygosity groups, largely limited to Generation -1 and 0.

Generation -1 mothers of SSDZ individuals were slightly older at their first birth (S5) as compared to mothers of MZ twins (β = 1.26), whereas mothers of XZ and multiples not in the FTC were significantly younger than mothers of MZ twins (XZ β = -0.68, other multiples β = - 2.41). This is consistent with prior research on DZ twinning and with other features of our sample (sampling bias and XZ twins, inclusion into the “multiples not in FTC” group”). Within Generation 0, OSDZ (β = -0.46) and XZ (β = -0.79) mothers were significantly younger at first birth as compared to MZ mothers. There were no differences for Generation 0 males for age at first birth. Nor were there zygosity differences in Generation 1for either males or females for age at first birth.

With respect to number of biological children in Generation 0 (Table S6), female SSDZ (IRR = 1.05) and OSDZ (IRR = 1.11) twins had more offspring than their female MZ counterparts, but XZ females had less children (IRR = 0.84). XZ males also had less children (IRR = 0.71) but there were no differences between MZ, SSDZ, and OSDZ males. Similarly, Generation 0 female SSDZ (OR = 0.81) and OSDZ (OR = 0.65) twins were less likely to be childless as compared to their MZ counterparts; and both XZ males (OR = 2.66) and females (OR = 2.15) were more likely to be childless (S7). SSDZ and OSDZ male twins did not differ from their MZ counterparts on likelihood of being childless. There were no zygosity differences in Generation 1 for number of biological children or being childless.

Female DZ twins were themselves more likely to have a multiple birth (Table S8) as compared to MZ female twins. Due to issues with statistical power, we were unable to evaluate whether the difference was observed specifically in SSDZ or OSDZ female twins, and if the effect differed between these two groups. We note the confidence intervals are particularly large, reflecting the rarity of the outcome, though the point estimates do follow the anticipated pattern (SSDZ and OSDZ both OR greater than 1).

#### Summary of Results Across Generations

Across three generations, fertility differences associated with twin status and zygosity were modest and largely confined to Generation 0. Female dizygotic twins, particularly those from opposite-sex pairs, exhibited modestly higher completed fertility than monozygotic twins and singletons. Twins were also more likely than singletons to have a multiple birth themselves, and this effect was specific to female DZ twins. However, these differences did not persist into the next generation. Fertility patterns among the children of twins were strikingly similar to those of the general population, providing no strong evidence for intergenerational transmission of zygosity-related fertility effects.

## Discussion

In this population-based register study linking the Finnish Twin Cohort with nationwide demographic data, we examined whether twin status and zygosity predict fertility patterns across multiple generations. Overall, twins in mid-20th-century Finland exhibited fertility outcomes that were highly similar to those of non-twins. Across nearly all indicators—including age at first birth, completed fertility, childlessness, and fertility in subsequent generations—observed differences were small in magnitude and unlikely to be demographically meaningful. The major exception being that twins were more likely than singletons to have a multiple birth, and this effect is specific to female dizygotic twins.

Female DZ twins had modestly higher completed fertility than MZ twin women, with a suggestive gradient indicating higher fertility among OSDZ twins compared with SSDZ twins. Although these differences were small in magnitude, their direction is consistent with established biological explanations for dizygotic twinning, particularly the heritable propensity toward hyperovulation in women (Hubers *et al*., 2025). This sex-specific pattern aligns with prior evidence that genetic influences on twinning act primarily through female reproductive physiology (Hubers *et al*., 2025; Mbarek *et al*., 2023) rather than through shared familial or social factors. We do not observe any evidence consistent with the opposing biological mechanisms proposed in Lummaa et al., 2007, consistent with other studies that do not observe this effect (Ahrenfeldt *et al*., 2015; Rose *et al*., 2002).

Importantly, these modest fertility advantages among female DZ twins did not extend to their offspring. The fertility of the children of twins was virtually identical to that of the children of singletons across all zygosity groups, both in the full sample and among those born before 1980. This finding suggests that while genetic factors related to DZ twinning may influence women’s own reproductive outcomes, they do not meaningfully shape fertility in subsequent generations. Taken together, these results indicate that any heritable component of twinning is specific and limited, rather than reflecting a broader transmission of high fertility across generations. The genetic component of twinning is highly polygenic (Hubers *et al*., 2025), and any genetic predisposition to hyperovulation may be diluted across generations due to Mendelian inheritance and the genetic contribution of the non-twin parent. Another factor may reduce parity in younger generations such that the genetic predisposition to twinning is not expressed when family sizes are typically one or two children in the most recent decades and fertility rates are low overall. Lastly, a limitation is the inability to confidently identify multiple births for Generation 1 individuals, limiting our ability to draw strong intergenerational conclusions about twin births.

In an evolutionary context, one reason that the “twinning advantage” doesn’t cascade down the generations is that evolution selects for fitness (the number of offspring who survive to reproduce), not just fecundity (the number of eggs or births; Orr, 2009). It is possible that the possible advantageous genetic fitness component of DZ twins (van Dongen *et al*., 2024) is counterbalanced by possible costs of twinning, for example high energetic demands of twin pregnancy, negative effects of prenatal exposure to sex-specific hormonal environments (Lummaa et al. 2007), and increased risks during labor. Consequently, while twinning may be selected in specific high-resource contexts (Lummaa *et al*., 1998), supported by the finding that mother’s physiological condition influences likelihood of twin pregnancy (Hubers *et al*., 2024), stabilizing selection seems to maintain singleton births as the dominant human reproductive strategy, preventing a broader transgenerational transmission of high fertility.

Our findings have important implications for twin research. Concerns have been raised that twins may differ systematically from the general population in reproductive or life-course outcomes, potentially limiting the generalizability of twin-based designs. The present results provide strong evidence against this concern for fertility-related traits, with the one exception of twin births by female dizygotic twins. Across a large, population-based cohort with complete multigenerational follow-up, twins and non-twins were remarkably similar in both timing and quantum of reproduction.

Our findings also inform us about potential selection effects and non-responder bias within the Finnish Twin Cohort data. Age at first birth was nearly identical between twins and singletons in both men and women. Small deviations were observed among twins classified as having unknown or uncertain zygosity, who tended to have earlier ages at first birth and lower completed fertility. However, this group consists of individuals who did not participate in survey-based zygosity determination or gave inconsistent responses to questions on zygosity in the surveys. They may therefore differ systematically from respondents of known zygosity. These patterns are consistent with response or selection bias rather than biological effects of twin status, and they do not undermine the overall similarity between twins and non-twins.

Strengths of this study include the use of nationwide register data, large sample sizes across multiple generations, and minimal loss to follow-up. Limitations include reliance on survey-based zygosity determination for some participants and the inability to directly observe the biological mechanisms underlying fertility differences or to measure genetic propensity to hyperovulation. Genotyping in later years has confirmed the validity of the survey-based method, which was first validated against blood groups (Kaprio *et al*., 2019; Sarna *et al*., 1978). Future research leveraging multigenerational genetic data, such as family-based polygenic scores, may provide a promising avenue for investigating these mechanisms. In addition, fertility patterns observed in mid-20th-century Finland may not generalize fully to contemporary populations with different social and reproductive norms, such as the widespread availability of assisted reproductive technologies like invitro fertilization, which have substantially altered the relationship between biological fecundity and realized fertility (Gissler and Tiitinen, 2001).

In conclusion, twin status does not meaningfully predict fertility outcomes for men or women, nor does it influence fertility in subsequent generations. Modest, sex-specific fertility differences among female DZ twins are consistent with known biological mechanisms of twinning but do not compromise the representativeness of twins as a population. These findings provide robust empirical support for the continued use of twin cohorts in studies of fertility and life-course outcomes.

## Author’s roles

SNP, SZ, and JK designed the study with input from MH on outcome operationalization. SNP and SZ completed data cleaning and analysis. SNP wrote the draft manuscript under the supervision of SZ. AL provided registry data access. All authors reviewed and agreed to the final version of the manuscript.

## Data Availability

The data underlying this article cannot be shared publicly due to the terms of data access specified in our ethical and permit approvals. To obtain comparable register data in Finland, you may contact Statistics Finland.

## Acknowledgements

We thank the Finnish population whose data makes our research possible and we appreciate the technical support from the FIONA platform IT.

## Ethics

Ethical approval was granted by the Finnish Institute for Health and Welfare, and data access is managed through the secure Statistics Finland FIONA platform. Data access was approved by Statistics Finland and FinData and subject to oversight by institutional ethics committees. All individual-level data were fully anonymized prior to analysis and accessed only through the secure FIONA environment. No personally identifiable data were used in any published output, and all analyses were conducted in accordance with approved research protocols.

