## supplemental tables for "Fertility rates across generations in twins and singletons: A total population study in Finland"

**Affiliations:**
¹ Institute for Molecular Medicine Finland, University of Helsinki, FI-00014 Helsinki, Finland

^2^ Minerva Foundation Institute for Medical Research, FI-00014 Helsinki, Finland

^3^ Institute of Criminology and Legal Policy, University of Helsinki, FI-00014 Helsinki, Finland

**Corresponding author:**
Stephanie Zellers

ORCID: 0000-0001-8927-3483

| **Table S1: Descriptives on Fertility of Children of Twins and Children of Singletons (Generation 1, Full Sample)** | | | | | | | | |
| --- | --- | --- | --- | --- | --- | --- | --- | --- |
|  | **N** | | **Mean Age at First Birth (SD)** | | **%Childless** | | **Mean N Children (SD)** | |
| **Twin Status** | **M** | **F** | **M** | **F** | **M** | **F** | **M** | **F** |
| Singletons | 625,991 | 598,785 | 29.2 (5.2) | 27.6  (5.2) | 40.8% | 30.8% | 1.30 (1.40) | 1.55 (1.42) |
| Twins | 19,490 | 18,784 | 29.3 (5.2) | 27.5  (5.1) | 40.0% | 29.9% | 1.32 (1.42) | 1.59 (1.45) |
| **Zygosity**  **Stratified** |  |  |  |  |  |  |  |  |
| MZ | 3,356 | 3,224 | 29.3 (5.2) | 27.4  (5.1) | 40.5% | 30.0% | 1.32 (1.38) | 1.61 (1.50) |
| SSDZ | 7,439 | 7,221 | 29.3 (5.2) | 27.6  (5.1) | 40.0% | 29.6% | 1.30 (1.41) | 1.59 (1.45) |
| OSDZ | 7,384 | 7,045 | 29.3 (5.2) | 27.6  (5.1) | 39.7% | 30.0% | 1.33 (1.45) | 1.58 (1.42) |
| XZ | 1,311 | 1,294 | 28.9 (5.3) | 26.8  (5.4) | 40.2% | 30.3% | 1.30 (1.39) | 1.59 (1.43) |

Note: M - males, F - females, MZ - monozygotic, SSDZ - same sex dizygotic, OSDZ - opposite sex dizygotic, XZ - unknown zygosity

| **Table S2: Twin vs. Singleton Differences in Age at First Birth for Full Gen 1 Sample** | | | | | |
| --- | --- | --- | --- | --- | --- |
| **Generation** | **Predictor** | **Males IRR (95% CI)** | **p-value** | **Females IRR (95% CI)** | **p-value** |
| Gen1 | Twin (yes vs no) | 0.08  (−0.03, 0.19) | 0.14 | −0.02  (−0.12, 0.08) | 0.69 |
|  | MZ | 0.17  (−0.09, 0.42) | 0.20 | −0.11  (−0.36, 0.13) | 0.37 |
|  | SSDZ | 0.11  (−0.06, 0.28) | 0.21 | 0.09  (−0.08, 0.25) | 0.30 |
|  | OSDZ | 0.09  (−0.08, 0.26) | 0.32 | 0.05  (−0.12, 0.21) | 0.60 |
|  | XZ | −0.31  (−0.71, 0.09) | 0.13 | **−0.75**  **(−1.13, −0.37)** | **1.3e-4** |

Note: Reference category is singletons, MZ - monozygotic, SSDZ - same sex dizygotic, OSDZ - opposite sex dizygotic, XZ - unknown zygosity; bolded cells are significant effects for alpha < 0.05.

| **Table S3: Twin vs. Singleton Differences in Total Number of Biological Children for Full Gen 1 Sample** | | | | | |
| --- | --- | --- | --- | --- | --- |
| **Generation** | **Predictor** | **Males IRR (95% CI)** | **p-value** | **Females IRR (95% CI)** | **p-value** |
| Gen1 | Twin (yes vs no) | 1.01  (0.99, 1.03) | 0.23 | **1.024**  **(1.008, 1.039)** | **0.002** |
|  | MZ | 1.01  (0.97, 1.05) | 0.73 | 1.02  (0.99, 1.06) | 0.21 |
|  | SSDZ | 1.00  (0.97, 1.03) | 0.99 | 1.03  (1.00, 1.05) | 0.05 |
|  | OSDZ | 1.02  (0.99, 1.05) | 0.14 | 1.02  (0.99, 1.05) | 0.08 |
|  | XZ | 1.02  (0.96, 1.09) | 0.54 | 1.02  (0.97, 1.08) | 0.40 |

Note: Reference category is singletons, MZ - monozygotic, SSDZ - same sex dizygotic, OSDZ - opposite sex dizygotic, XZ - unknown zygosity; bolded cells are significant effects for alpha < 0.05.

| **Table S4: Twin vs. Singleton Differences in Likelihood of Being Childless for Full Gen 1 Sample** | | | | | |
| --- | --- | --- | --- | --- | --- |
| **Generation** | **Predictor** | **Males IRR (95% CI)** | **p-value** | **Females IRR (95% CI)** | **p-value** |
| Gen1 | Twin (yes vs no) | 0.97  (0.94, 1.00) | 0.07 | **0.96**  **(0.92, 0.99)** | **0.02** |
|  | MZ | 1.00  (0.92, 1.08) | 0.97 | 0.97  (0.89, 1.06) | 0.45 |
|  | SSDZ | 0.97  (0.92, 1.02) | 0.23 | **0.94**  **(0.88, 0.99)** | **0.04** |
|  | OSDZ | 0.96  (0.91, 1.01) | 0.12 | 0.96  (0.91, 1.03) | 0.23 |
|  | XZ | 0.97  (0.86, 1.10) | 0.66 | 0.99  (0.86, 1.14) | 0.90 |

Note: Reference category is singletons, MZ - monozygotic, SSDZ - same sex dizygotic, OSDZ - opposite sex dizygotic, XZ - unknown zygosity; bolded cells are significant effects for alpha < 0.05.

| **Table S5: Twin-Only Zygosity Contrasts for Age at First Birth** | | | | | |
| --- | --- | --- | --- | --- | --- |
| **Generation** | **Predictor** | **Males β (95% CI)** | **p-value** | **Females β (95% CI)** | **p-value** |
| Gen -1 | DZ | — | — | **0.73**  **(0.38, 1.08)** | **4.0e-5** |
|  | SSDZ | — | — | **1.26**  **(0.87, 1.64)** | **1.10e-10** |
|  | OSDZ | — | — | 0.25  (-0.13, 0.63) | 0.19 |
|  | XZ | — | — | **-0.68**  **(-1.13, -0.23)** | **0.003** |
|  | Twins/Multiples not in FTC | — | — | **-2.41**  **(-2.75, -2.07)** | **<2e-16** |
| Gen 0 | DZ | 0.20  (-0.13, 0.53) | 0.24 | **-0.32**  **(-0.62, -0.02)** | **0.04** |
|  | SSDZ | 0.20  (-0.16, 0.56) | 0.28 | -0.15  (-0.47, 0.18) | 0.38 |
|  | OSDZ | 0.20  (-0.16, 0.56) | 0.28 | **-0.46**  **(-0.77, -0.14)** | **0.005** |
|  | XZ | 0.18  (-0.33, 0.69) | 0.50 | **-0.79**  **(-1.30, -0.28)** | **0.002** |
| Gen 1 Born by 1980 | DZ | -0.12  (-0.48, 0.24) | 0.51 | 0.27  (-0.08, 0.61) | 0.13 |
|  | SSDZ | -0.10  (-0.49, 0.28) | 0.61 | 0.26  (-0.11, 0.63) | 0.17 |
|  | OSDZ | -0.14  (-0.53, 0.25) | 0.48 | 0.27  (-0.10, 0.65) | 0.16 |
|  | XZ | -0.46  (-1.07, 0.14) | 0.13 | -0.45  (-1.02, 0.11) | 0.12 |
| All Gen 1 | DZ | -0.07  (-0.34, 0.20) | 0.61 | 0.16  (-0.10, 0.42) | 0.23 |
|  | SSDZ | -0.06  (-0.36, 0.24) | 0.69 | 0.17  (-0.11, 0.46) | 0.23 |
|  | OSDZ | -0.08  (-0.38, -0.21) | 0.59 | 0.15  (-0.14, .0.43) | 0.31 |
|  | XZ | -0.44  (-0.90, 0.02) | 0.06 | **-0.65**  **(-1.08, -0.20)** | **0.004** |

Note: Reference category is MZ, MZ - monozygotic, DZ - all dizygotic (same and opposite sex), SSDZ - same sex dizygotic, OSDZ - opposite sex dizygotic, XZ - unknown zygosity; bolded cells are significant effects for alpha < 0.05.

| **Table S6: Twin-Only Zygosity Contrasts for Total Number of Biological Children** | | | | | |
| --- | --- | --- | --- | --- | --- |
| **Generation** | **Predictor** | **Males IRR (95% CI)** | **p-value** | **Females IRR (95% CI)** | **p-value** |
| Gen -1 | DZ | — | — | 1.03  (1.00, 1.06) | 0.09 |
|  | SSDZ | — | — | 1.00  (0.96, 1.03) | 0.88 |
|  | OSDZ | — | — | **1.06**  **(1.02, 1.10)** | **0.002** |
|  | XZ | — | — | **1.14**  **(1.09, 1.18)** | **7.9e-10** |
|  | Twins/Multiples not in FTC | — | — | **1.40**  **(1.36, 1.45)** | **<2e-16** |
| Gen 0 | DZ | 1.01  (0.96, 1.06) | 0.64 | **1.08**  **(1.04, 1.13)** | **1.6e-4** |
|  | SSDZ | 1.01  (0.96, 1.06) | 0.81 | **1.05**  **(1.00, 1.10)** | **0.03** |
|  | OSDZ | 1.02  (0.96, 1.07) | 0.55 | **1.11**  **(1.06, 1.16)** | **2.6e-6** |
|  | XZ | **0.71**  **(0.66, 0.77)** | **<2e-16** | **0.84**  **(0.79, 0.90)** | **1.3e-6** |
| Gen 1 Born by 1980 | DZ | 1,03  (0.98, 1.08) | 0.20 | 1.00  (0.05, 1.04) | 0.83 |
|  | SSDZ | 1.05  (0.96, 1.07) | 0.55 | 0.99  (0.95, 1.04) | 0.75 |
|  | OSDZ | 1.02  (0.99, 1.11) | 0.08 | 1.00  (0.95, 1.05) | 0.95 |
|  | XZ | 1.05  (0.94, 1.10) | 0.67 | 0.99  (0.92, 1.06) | 0.73 |
| All Gen 1 | DZ | 1.00  (0.96, 1.05) | 0.94 | 1.00  (0.96, 1.04) | 0.97 |
|  | SSDZ | 0.99  (0.94, 1.04) | 0.72 | 1.00  (0.96 1.05) | 0.85 |
|  | OSDZ | 1.01  (0.96, 1.06) | 0.61 | 1.00  (0.96, 1.04) | 0.97 |
|  | XZ | 1.01  (0.94, 1.09) | 0.77 | 1.00  (0.94, 1.07) | 0.93 |

Note: Reference category is MZ, MZ - monozygotic, DZ - all dizygotic (same and opposite sex), SSDZ - same sex dizygotic, OSDZ - opposite sex dizygotic, XZ - unknown zygosity; bolded cells are significant effects for alpha < 0.05.

| **Table S7: Twin-Only Zygosity Contrasts for Likelihood of Being Childless** | | | | | |
| --- | --- | --- | --- | --- | --- |
| **Generation** | **Predictor** | **Males OR (95% CI)** | **p-value** | **Females OR (95% CI)** | **p-value** |
| Gen 0 | DZ | 1.04  (0.91, 1.19) | 0.57 | **0.72**  **(0.62, 0.85)** | **6.9e-5** |
|  | SSDZ | 1.01  (0.87, 1.17) | 0.88 | **0.81**  **(0.68, 0.97)** | **0.02** |
|  | OSDZ | 1.07  (0.92, 1.24) | 0.38 | **0.65**  **(0.55, 0.78)** | **2.0e-6** |
|  | XZ | **2.66**  **(2.22, 3.19)** | **<2e-16** | **2.15**  **(1.69, 2.73)** | **4.7e-10** |
| Gen 1 Born by 1980 | DZ | 0.91  (0.80, 1.02) | 0.11 | 1.02  (0.88, 1.18) | 0.81 |
|  | SSDZ | 0.93  (0.81, 1.06) | 0.27 | 1.02  (0.87, 1.20) | 0.81 |
|  | OSDZ | 0.88  (0.77, 1.01) | 0.07 | 1.02  (0.86, 1.19) | 0.85 |
|  | XZ | 1.01  (0.82, 1.24) | 0.93 | 1.04  (0.81, 1.32) | 0.77 |
| All Gen 1 | DZ | 0.97  (0.89, 1.05) | 0.43 | 0.99  (0.90, 1.08) | 0.77 |
|  | SSDZ | 0.97  (0.88, 1.07) | 0.54 | 0.97  (0.88, 1.08) | 0.62 |
|  | OSDZ | 0.96  (0.88, 1.06) | 0.41 | 1.00  (0.90, 1.11) | 0.97 |
|  | XZ | 0.98  (0.84, 1.13) | 0.73 | 1.02  (0,87, 1.20) | 0.77 |

Note: Reference category is MZ, MZ - monozygotic, DZ - all dizygotic (same and opposite sex), SSDZ - same sex dizygotic, OSDZ - opposite sex dizygotic, XZ - unknown zygosity; bolded cells are significant effects for alpha < 0.05.

| **Table S8: Twin-Only Zygosity Contrasts for Likelihood of Having Twin Offspring** | | | | | |
| --- | --- | --- | --- | --- | --- |
| Generation | Predictor | Males OR (95% CI) | p-value | Females OR (95% CI) | p-value |
| Gen 0 | DZ | 1.43  (0.34, 6.01) | 0.62 | **2.36**  **(1.25, 4.46)** | **0.01** |
|  | SSDZ | 1.08  (0.22, 5.26) | 0.92 | 1.34  (0.30, 5.99) | 0.70 |
|  | OSDZ | 1.82  (0.40, 8.18) | 0.44 | 2.62  (0.64, 10.62) | 0.18 |
|  | XZ | 0.94  (0.11, 7.73) | 0.95 | 1.05  (0.11, 10.01) | 0.97 |

Note: Reference category is MZ, MZ - monozygotic, DZ - all dizygotic (same and opposite sex), SSDZ - same sex dizygotic, OSDZ - opposite sex dizygotic, XZ - unknown zygosity; bolded cells are significant effects for alpha < 0.05.
